# A Systematic Meta-Analysis of CT Features of COVID-19: Lessons from Radiology

**DOI:** 10.1101/2020.04.04.20052241

**Authors:** Vasantha Kumar Venugopal, Vidur Mahajan, Sriram Rajan, Vikash Agarwal, Ruchika Rajan, Salsabeel Syed, Harsh Mahajan

## Abstract

Several studies have been published in the past few months describing the CT features of Coronavirus Disease 2019 (COVID-19). There is a great degree of heterogeneity in the study designs, lesion descriptors used and conclusions derived. In our systematic analysis and meta-review, we have attempted to homogenize the reported features and provide a comprehensive view of the disease pattern and progression in different clinical stages. After an extensive literature search, we short-listed and reviewed 49 studies including over 4145 patients with 3615 RT-PCR positive cases of COVID-19 disease. We have found that there is a good agreement among these studies that diffuse bilateral ground-glass opacities (GGOs) is the most common finding at all stages of the disease followed by consolidations and mixed density lesions. 78% of patients with RT-PCR confirmed COVID-19 infections had either ground-glass opacities, consolidation or both. Inter-lobular septal thickening was also found to be a common feature in many patients in advanced stages. The progression of these initial patchy GGO’s and consolidations to diffuse lesions with septal thickening, air bronchograms in the advanced stages, to either diffuse “white-out” lungs needing ICU admissions or finally resolving completely without or with residual fibrotic strips was also found to be congruent among multiple studies. Prominent juxta- lesional pulmonary vessels, pleural effusion and lymphadenopathy in RT-PCR proven cases were found to have poor clinical prognosis. Additionally, we noted wide variation in terminology used to describe lesions across studies and suggest the use of standardized lexicons to describe findings related to diseases of vital importance.

## 2. Introduction

Severe Acute Respiratory Syndrome Coronavirus 2 (SARS-CoV-2), or simply COVID-19 has spread to pandemic proportions across the world. As of 30^th^ March 2020, the disease had affected nearly 700,000 people causing 33,000 deaths, with the United States becoming the latest epicenter with more than 100,000 cases and Italy reporting the highest number of deaths - over 10,000 people. Several thousands of pieces of the scientific literature have been published over the past few months on the clinical manifestations, epidemiology, molecular biology, imaging features and lab diagnosis of COVID-19. Amongst these, many studies reported the radiological findings of the disease in various stages and some even compared the performance of medical imaging to Real-Time Polymerase Chain Reaction (RT-PCR) for initial diagnosis. Several imaging societies have released recommendations for the use of imaging in the diagnosis of COVID-19 **(1)**. This has led to a plethora of disjointed information available, with different qualitative descriptors making it difficult for a practicing radiologist to keep abreast of scientific developments. We have performed this systematic review & meta-analysis of the literature available on the PubMed database reporting the manifestations of COVID-19 disease on Computed Tomography (CT) scans, synthesized several data points from these studies, and attempted to present a comprehensive analysis of all the reported CT findings of the disease.

## 3. Methods and Materials

We performed a systematic search of the literature on the PubMed database initially on March 10, 2020, using the terms “novel coronavirus”, “nCov”, “COVID-19” and “CT”, “Computed Tomography” as keywords using the advanced search function. The search was repeated on March 26, 2020, to include new reports. The studies were analyzed for their quality and appropriateness. Duplicate studies were removed. Additional studies were also identified from the references cited in the shortlisted studies.

Shortlisted studies were distributed amongst five radiologists (^1^SR, RR, SS, VA, HM), who systemically analyzed each to quantify the imaging features on CT scan studies described in the studies. Other relevant data categories including patient demographics, type of study, location of the study hospitals, RT-PCR results were also recorded along with disease progression information and clinical outcomes, wherever available.

## 4. Results

We found a total of 677 studies using the Advanced Search function on PubMed. 59 studies were finally shortlisted after quality assessment. After the removal of 10 studies due to lack of sufficient case-level information, there were 49 studies including 16 case series and reports, and 33 studies with reports of 9 or more patients (Figure 1). Together, these studies included a total of 4145 patients with 3615 (87%) of them being RT-PCR positive cases for COVID-19 disease. 530 (13%) patients were negative for COVID-19 disease on RT-PCR testing. There were two studies exclusively done on pediatric patients which included 23 pediatric patients. The other studies together had 10 children in their study sets. The gender distribution information of the CT scans was not clearly described in two studies. Among the remaining patients, there were 1,512 males and 1,579 females in the age range of 16 to 92 years.

**Figure 1.**
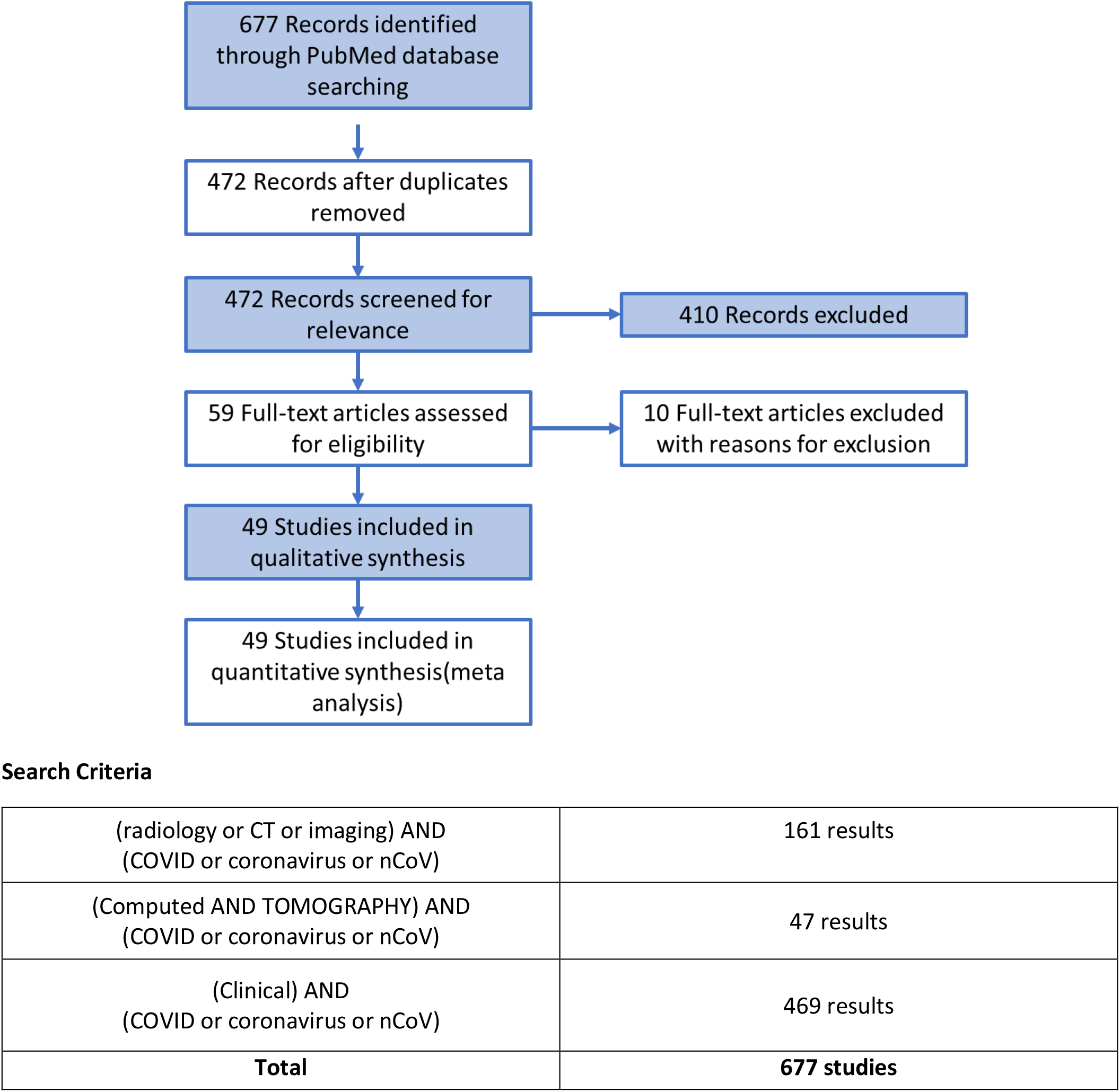
Study method, step-wise results and Search Criteria. <u>PRISMA (Preferred Reporting Items for Systematic Reviews and Meta-Analyses)</u>

**Figure 2.**
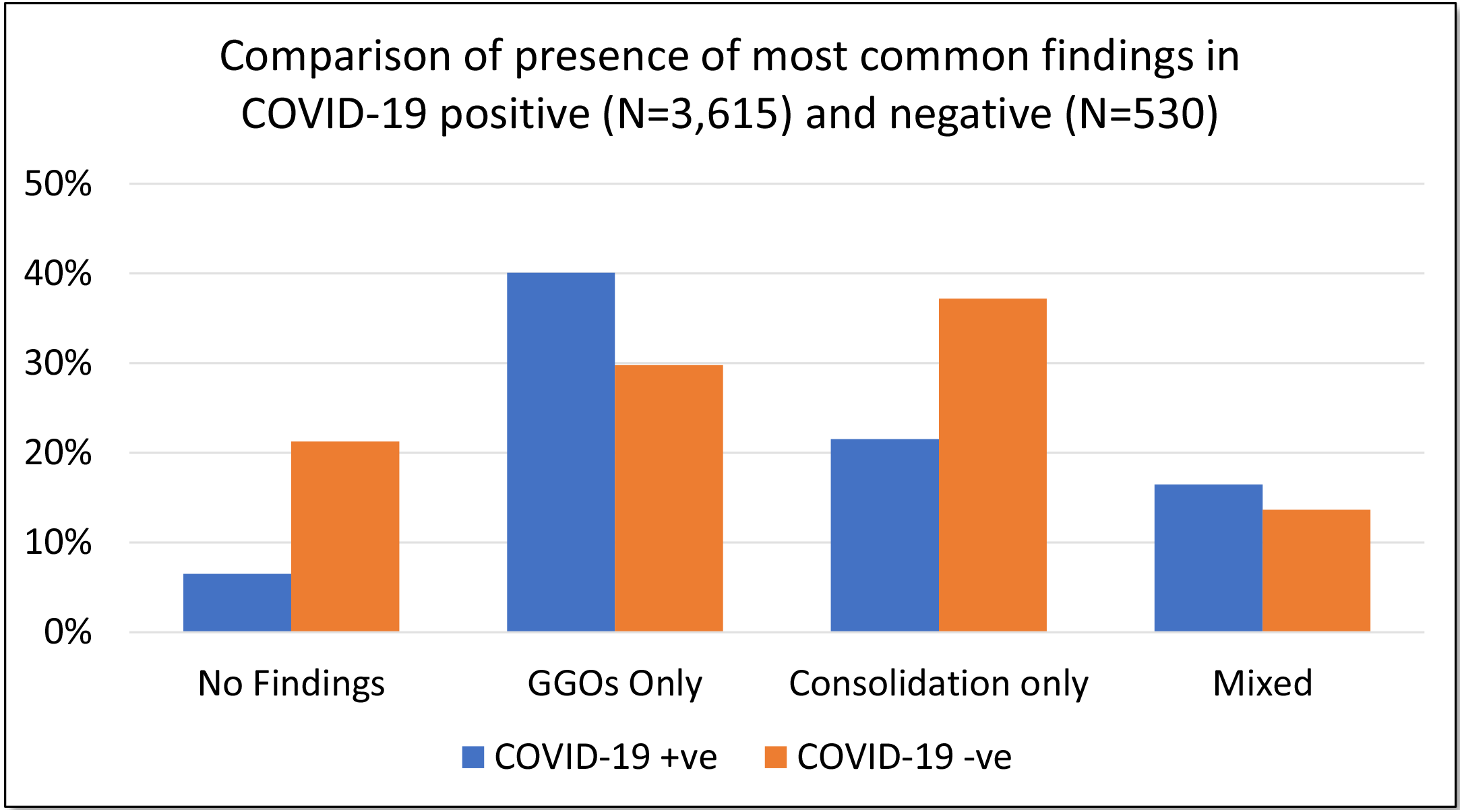
A bar chart displaying comparison (in %) of the most common CT features between RT-PCR positive (N=3,615) COVID-19 cases and RT-PCR negative (N=530) cases. Ground Glass Opacities (GGOs) and Mixed lesions were more common in positive cases and consolidation was seen more prevent in negative cases.

Our review of 49 studies included 46 studies from china, one report each from South Korea, Italy and Germany. Two studies by Ai et al.and Guan et al.**(4,28)**, published in late February, included a large series of patients (>1000 patients). The earliest report was a retrospective analysis of 99 cases by Cheng et al. in late January. All the studies, including the ones which were excluded, are shown in Tables 1 & 2.

**Table 1.** All Included Studies, Sorted by Date of Publication.

|  | Title | Type | Author | Date | Origin | Journal |
| --- | --- | --- | --- | --- | --- | --- |
| 1 | CT Imaging of the 2019 Novel Coronavirus (2019-nCoV) Pneumonia | Case Report | Lei et al. | 31-01-2020 | China | Radiology |
| 2 | Chest Imaging Appearance of COVID-19 Infection | Case Report | Kong et al. | 01-02-2020 | China | Radiology: Cardiothoracic Imaging |
| 3 | Imaging Profile of the COVID-19 Infection: Radiologic Findings and Literature Review | Retrospective | Ng et al. | 01-02-2020 | China | Radiology: Cardiothoracic Imaging |
| 4 | CT Imaging Features of 2019 Novel Coronavirus (2019-nCoV) | Retrospective | Chung et al. | 04-02-2020 | China | Radiology |
| 5 | Emerging Coronavirus 2019-nCoV Pneumonia | Retrospective | Song et al. | 06-02-2020 | China | Radiology |
| 6 | A rapid advice guideline for the diagnosis and treatment of 2019 novel coronavirus (2019-nCoV) infected pneumonia (standard version) | Guideline | Jin et al. | 06-02-2020 | China | Military Medical Research |
| 7 | CT Manifestations of Two Cases of 2019 Novel Coronavirus (2019-nCoV) Pneumonia | Case Report | Fang et al. | 07-02-2020 | China | Radiology |
| 8 | Epidemiologic and Clinical Characteristics of Novel Coronavirus Infections Involving 13 Patients Outside Wuhan, China | Case Report | Chang et al. | 07-02-2020 | China | JAMA |
| 9 | Clinical and biochemical indexes from 2019-nCoV infected patients linked to viral loads and lung injury | Retrospective | Liu et al. | 12-07-1905 | China | Science China. Life Sciences |
| 10 | Chest CT for Typical 2019-nCoV Pneumonia: Relationship to Negative RT-PCR Testing | Case Report | Xie et al. | 12-02-2020 | China | Radiology |
| 11 | Time Course of Lung Changes On Chest CT During Recovery From 2019 Novel Coronavirus (COVID-19) Pneumonia | Retrospective | Pan et al. | 13-02-2020 | China | Radiology |
| 12 | Initial CT Findings and Temporal Changes in Patients With the Novel Coronavirus Pneumonia (2019-nCoV): A Study of 63 Patients in Wuhan, China | Retrospective | Pan et al. | 13-02-2020 | China | European Radiology |
| 13 | A familial cluster of pneumonia associated with the 2019 novel coronavirus indicating person-to-person transmission: a study of a family cluster | Case Report | Chan et al. | 15-02-2020 | China | The Lancet |
| 14 | Sensitivity of Chest CT for COVID-19: Comparison to RT-PCR | Retrospective | Yang et al. | 19-02-2020 | China | Radiology |
| 15 | Chest CT Findings in Coronavirus Disease-19 (COVID-19): Relationship to Duration of Infection | Retrospective | Bernheim et al. | 20-02-2020 | China | Radiology |
| 16 | Clinical characteristics and imaging manifestations of the 2019 novel coronavirus disease (COVID-19): A multi-center study in Wenzhou city, Zhejiang, China | Retrospective | Yang et al. | 21-02-2020 | China | Journal of Infection |
| 17 | Asymptomatic novel coronavirus pneumonia patient outside Wuhan: The value of CT images in the course of the disease | Case Report | Lin et al. | 01-07-2020 | China | Clinical Imaging |
| 18 | Radiological findings from 81 patients with COVID-19 pneumonia in Wuhan, China: a descriptive study | Retrospective | Shi et al. | 01-04-2020 | China | The Lancet Infectious Diseases |
| 19 | Clinical and computed tomographic imaging features of novel coronavirus pneumonia caused by SARS-CoV-2 | Retrospective | Xu et al. | 25-02-2020 | China | The Journal of Infection |
| 20 | 2019 Novel Coronavirus (COVID-19) Pneumonia: Serial Computed Tomography Findings | Case Report | Wei et al. | 12-07-1905 | Korea | Korean Journal of Radiology |
| 21 | Correlation of Chest CT and RT-PCR Testing in Coronavirus Disease 2019 (COVID-19) in China: A Report of 1014 Cases | Retrospective | Ai et al. | 26-02-2020 | China | Radiology |
| 22 | 2019 Novel Coronavirus (COVID-19) Pneumonia with Hemoptysis as the Initial Symptom: CT and Clinical Features | Case Report | Shi F et al | 26-02-2020 | China | Korean Journal of Radiology |
| 23 | 2019 Novel Coronavirus (COVID-19) Pneumonia with Hemoptysis as the Initial Symptom: CT and Clinical Features | Case Report | F Shi et al. | 26-02-2020 | China | Korean Journal of Radiology |
| 24 | 2019-novel Coronavirus severe adult respiratory distress syndrome in two cases in Italy: An uncommon radiological presentation | Case Report | Albarelli et al. | 01-04-2020 | Italy | International Journal of Infectious Diseases |
| 25 | Chest Radiographic and CT Findings of the 2019 Novel Coronavirus Disease (COVID-19): Analysis of Nine Patients Treated in Korea | Retrospective | Yoon et al. | 12-07-1905 | Korea | Korean Journal of Radiology |
| 26 | Imaging and clinical features of patients with 2019 novel coronavirus SARS-CoV-2 | Retrospective | Xu et al. | 28-02-2020 | China | European Journal of Nuclear Medicine and Molecular Imaging |
| 27 | Clinical Characteristics of Coronavirus Disease 2019 in China | Retrospective | Guan et al. | 28-02-2020 | China | NEJM |
| 28 | Clinical Characteristics of Imported Cases of COVID-19 in Jiangsu Province: A Multicenter Descriptive Study | Retrospective | Wu et al. | 29-02-2020 | China | Clinical Infectious Diseases |
| 29 | Relation Between Chest CT Findings and Clinical Conditions of Coronavirus Disease (COVID-19) Pneumonia: A Multicenter Study | Retrospective | Zhao et al. | 03-03-2020 | China | American Journal of Roentgenology |
| 30 | Coronavirus Disease 2019 (COVID-19): Role of Chest CT in Diagnosis and Management | Retrospective | Li et al. | 04-03-2020 | China | American Journal of Roentgenology |
| 31 | CT Imaging and Differential Diagnosis of COVID-19 | Case Report | Dai et al. | 04-03-2020 | China | Canadian Association of Radiologists Journal |
| 32 | Clinical and CT Features in Pediatric Patients With COVID-19 Infection: Different Points From Adults | Retrospective | Xia et al. | 00-01-1900 | China | Pediatric Pulmonology |
| 33 | CT Features of Coronavirus Disease 2019 (COVID-19) Pneumonia in 62 Patients in Wuhan, China | Retrospective | Zhou et al. | 05-03-2020 | China | American Journal of Roentgenology |
| 34 | Chest CT Findings in Patients with Corona Virus Disease 2019 and its Relationship with Clinical Features | Retrospective | Wu et al. | 06-03-2020 | China | Investigative Radiology |
| 35 | The Clinical and Chest CT Features Associated with Severe and Critical COVID-19 Pneumonia | Retrospective | Li et al. | 06-03-2020 | China | Investigative Radiology |
| 36 | Clinical and High-Resolution CT Features of the COVID-19 Infection: Comparison of the Initial and Follow-up Changes | Retrospective | Xiong et al. | 06-03-2020 | China | Investigative Radiology |
| 37 | FDG PET/CT of COVID-19 | Case Report | Zou et al. | 06-03-2020 | China | Radiology |
| 38 | Clinical characteristics and intrauterine vertical transmission potential of COVID-19 infection in nine pregnant women: a retrospective review of medical records | Retrospective | Chen et al. | 07-03-2020 | China | The Lancet |
| 39 | CT manifestations of coronavirus disease-2019: A retrospective analysis of 73 cases by disease severity | Retrospective | Liu KC et al. | 01-05-2020 | China | European Journal of Radiology |
| 40 | Initial clinical features of suspected Coronavirus Disease 2019 in two emergency departments outside of Hubei, China | Retrospective | Zhu et al. | 00-01-1900 | China | Journal of Medical Virology |
| 41 | Clinical Features and Chest CT Manifestations of Coronavirus Disease 2019 (COVID-19) in a Single-Center Study in Shanghai, China | Retrospective | Cheng et al. | 14-03-2020 | China | American Journal of Roentgenology |
| 42 | Clinical features of pediatric patients with COVID-19: a report of two family cluster cases | Case Report | Li-Na Ji et al. | 16-03-2020 | China | World Journal of Pediatrics |
| 43 | Successful recovery of COVID-19 pneumonia in a renal transplant recipient with long-term immunosuppression | Case Report | Zhu et al. | 00-01-1900 | China | American Journal of Transplantation |
| 44 | Early Clinical and CT Manifestations of Coronavirus Disease 2019 (COVID-19) Pneumonia | Retrospective | Han et al. | 17-03-2020 | China | American Journal of Roentgenology |
| 45 | Clinical Characteristics of 138 Hospitalized Patients With 2019 Novel Coronavirus-Infected Pneumonia in Wuhan, China | Retrospective | Wang et al. | 17-03-2020 | China | JAMA |
| 46 | Clinical progression of patients with COVID-19 in Shanghai, China | Retrospective | Chen at al | 19-03-2020 | China | Journal of Infection |
| 47 | CT appearance of severe, laboratory-proven coronavirus disease 2019 (COVID-19) in a Caucasian patient in Berlin, Germany | Case Report | Gross et al. | 19-03-2020 | Germany | RoFo |
| 48 | Temporal Changes of CT Findings in 90 Patients with COVID-19 Pneumonia: A Longitudinal Study | Retrospective | Wang et al. | 19-03-2020 | China | Radiology |
| 49 | High-resolution computed tomography manifestations of COVID-19 infections in patients of different ages | Retrospective | Chen et al. | 24-03-2020 | China | European Journal of Radiology |

**Table 2.** – All Excluded Studies, Sorted by Date of Publication.

|  | **Title** | **Type** | **Author** | **Date** | **Journal** | **Reason for Exclusion** |
| --- | --- | --- | --- | --- | --- | --- |
| **1** | Clinical features of patients infected with 2019 novel coronavirus in Wuhan, China | Retrospective | Huang et al. | 15-02-2020 | The Lancet | Adequate data not provided |
| **2** | 18F-FDG PET/CT findings of COVID-19: a series of four highly suspected cases | Case Report | Qin et al. | 22-02-2020 | European Journal of Nuclear Medicine and Molecular Imaging | RT-PCR status unknown for patients |
| **3** | COVID-19 pneumonia: what has CT taught us? | Perspective | Lee et al. | 24-02-2020 | Lancet Infectious Diseases | Perspective article |
| **4** | Radiology Perspective of Coronavirus Disease 2019 (COVID-19): Lessons From Severe Acute Respiratory Syndrome and Middle East Respiratory Syndrome | Perspective | Hesseiny et al. | 28-02-2020 | American Journal of Roentgenology | Perspective article |
| **5** | Study on the Distribution Characteristics and Signs of CT of 130 Cases of New Coronavirus Pneumonia | Retrospective | Wu et al. | 03-03-2020 | Chinese Journal of Tuberculosis & Respiratory Diseases | Paper in Chinese language |
| **6** | Patients with RT-PCR Confirmed COVID-19 and Normal Chest CT | Perspective | Yang et al. | 06-03-2020 | Radiology | Perspective article |
| **7** | Coronavirus Disease 2019 (COVID-19): A Systematic Review of Imaging Findings in 919 Patients | Meta-analysis | Salehi et al. | 14-03-2020 | American Journal of Roentgenology | Meta-analysis article |
| **8** | Artificial Intelligence Distinguishes COVID-19 from Community Acquired Pneumonia on Chest CT | Retrospective | Li et al. | 19-03-2020 | Radiology | CT findings not mentioned |
| **9** | The Progression of Computed Tomographic (CT) Images in Patients With Coronavirus Disease (COVID-19) Pneumonia: The CT Progression of COVID-19 Pneumonia | Retrospective | Pinggui Lei | 20-03-2020 | Journal of Infection | Adequate data not provided |
| **10** | Clinical characteristics of novel coronavirus cases in tertiary hospitals in Hubei Province | Retrospective | Liu et al. | 23-03-2020 | Chinese Medical Journal | Adequate data not provided |

### Imaging features on CT

*Note: A supplementary document which describes the number of cases of each finding, along with the nomenclature dictionary used, is available along with this manuscript*.

The lesions on CT scans of COVID-19 patients have been reported primarily in the following dimensions - distribution, number, shape, and density. The most frequent descriptor for the distribution of the lesions was the presence of bilateral lung lesions. Of the total 3514 scans of RT PCR positive cases of COVID-19 with information of laterality, 2364 (67%) had been reported to have bilateral involvement. Almost all the studies with information on the axial spatial distribution, 27 studies, agreed that the predominant pattern is peripheral and subpleural distribution. The predominant lobar distribution information was available in 19 studies, with 12 studies reporting right lower lobe as the most commonly involved lobe followed by the left lower lobe in 11 studies. The percentage of cases with right lower lobe involvement ranged from 27 to 79% **(2,3)**.

Among a total of 3,615 RTPCR positive patients included in this study, 235 (7%) patients had no findings on CT scans, most of the RT-PCR confirmed cases had the CT scans either in the initial stages of the onset of symptoms or before the onset of symptoms **(4,5)**.

There is universal agreement among all studies that ground glass opacification (GGO) is the most common abnormality regardless of the stage of the disease. GGO without consolidation is the most frequent finding in our review and it was seen in 1449 patients (40%). It was followed by consolidations in 778 patients (22%). Ground glass opacities with consolidation (mixed presentation) were seen 596 patients (16%). In these studies, 78% of patients with RT-PCR confirmed COVID-19 infections had either ground-glass opacities, consolidation or both. Several studies have reported GGOs in the early phase of the disease progressing to consolidations and later in the more severe phase a mixed picture **(5-7)**.

Interlobular septal thickening was seen in a total of 307 cases (14%), commonly in the advanced stages of the disease process (> 1 week after symptom onset) **(8,9)**. They are often seen along with ground glass opacities. The appearance of septal thickening is associated with the progression of the clinical severity of the disease.

The prominence of pulmonary vessels in relation to the lesion is reported in 228 patients overall. The only report from Italy included in this review reported that the perilesional pulmonary vessel enlargement in could an early predictor of lung impairment **(10)**. Similar appearances were previously reported in some reports from china under different terminologies like vascular engorgement **(11)**, vascular thickening **(3)**, microvascular dilatation **(12)**, and vascular enlargement **(13)**. This finding was seen in 45% to 82% of the cases in studies that reported them, all of them in radiology journals.

GGO with intralobular septal thickening (also known as the crazy-paving pattern) was reported in a total of 198 patients (5%), slightly higher frequency compared to fibrotic changes like linear atelectasis or fibrotic stripes seen in 176 patients (5%). Intralobular septal thickening or reticulations were reported in a total of 149 patients (4%), and all the patients had associated ground-glass opacities **(13,14)**.

These findings, however, were seen in different disease courses, the former often seen in advanced disease stages needing ICU admission, usually within two weeks of the onset of the symptoms whereas the latter was observed during the dissipation stage of the disease usually at two to three weeks after the onset of symptoms **(5)**.

Specific signs like halo sign and reverse halo sign are infrequently described in these reviewed studies. ‘Halo sign’ was seen in 88 cases in total, from three studies **(3, 11 & 26)**. 69 cases with the ‘Halo Sign’ were reported in a study of 108 patients by Han et al.**(3)**. Li et al.observed halo sign around nodules in 9 (17.6%) patients out of a series of 51 patients. Xia et al.in their pediatric series of 20 patients noted halo sign in 10 patients (50%) around consolidations **(26)**. ‘Halo sign’ refers to an area of GGO opacity around a nodule or consolidation. Similarly, “reversed halo sign”, described as a focal rounded area of ground-glass opacity surrounded by a more or less complete ring of consolidation **(Figure 5)** were reported only in four studies **(11, 21, 30 & 31)**. Out of the 240 patients included in these studies, CT scans of only 7 patients demonstrated this sign. Some studies may have included these findings as a part of mixed ground-glass opacities. It may be pertinent to study the frequency of these findings in various stages of the disease. In this sense, it is surprising that the recently released consensus statement by the Radiological Society of North America endorsed by the Society of Thoracic Radiology and the American College of Radiology recommends reporting the “Reverse Halo” sign as one of the typical findings of COVID-19 **(1)**. The referenced study by Bernheim et al. for this recommendation reported reverse halo sign only in two cases out of 121 patients.

**Figure 5.**
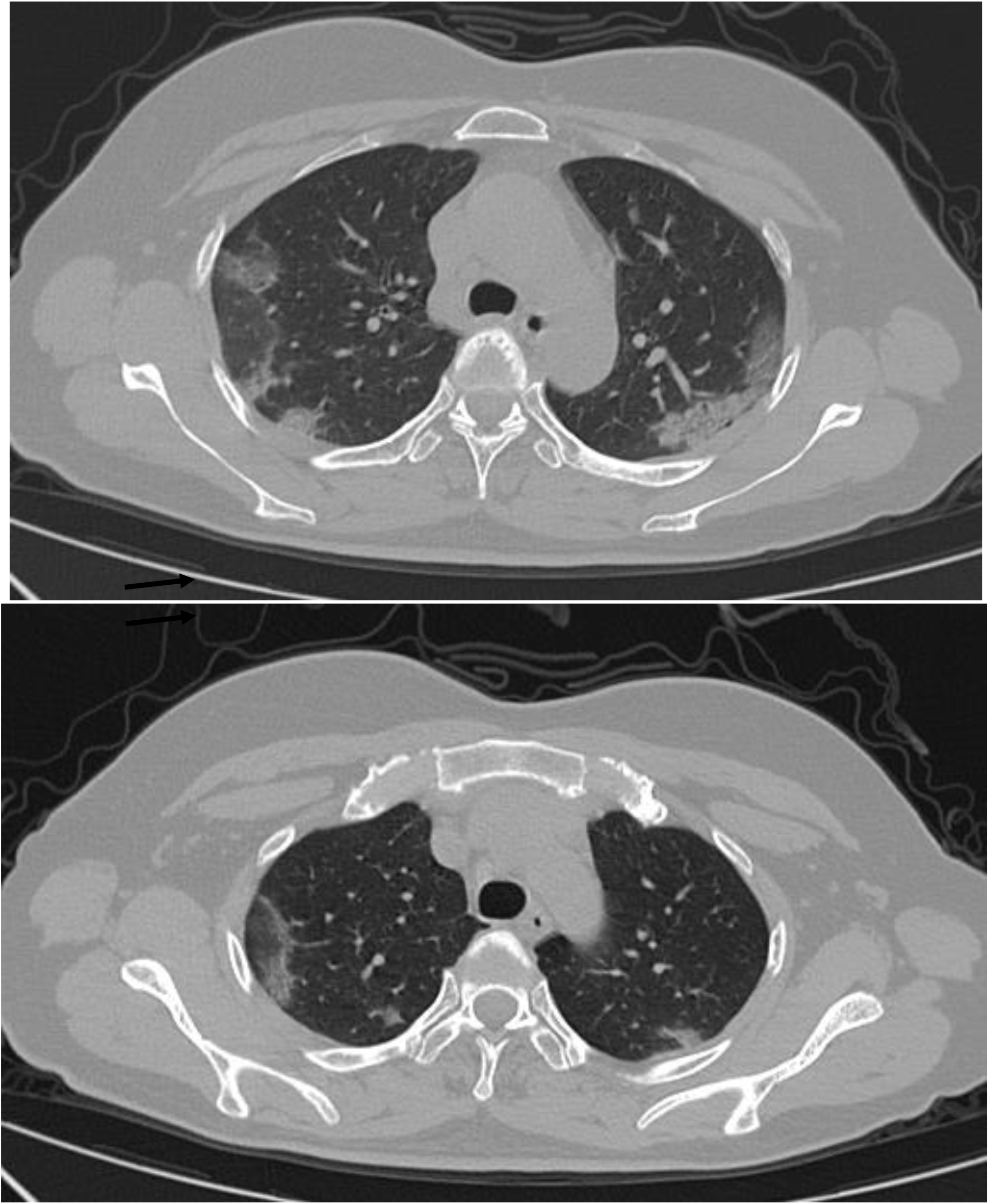
CT findings in a case of COVID-19 demonstrate bilateral peripheral ground-glass opacities with reticulation. There is also “reversed halo” appearance in the right upper lobe lesion (image courtesy: www.coronacases.org)

Air bronchogram, an essential sign of lobar pneumonia, were reported in 262 patients (10%). This is a pattern of air-filled (low-attenuation) bronchi on a background of opaque (high-attenuation) airless lung **(15)**. This was also seen more frequently in advanced stages of the disease, typically in the second week after onset of symptoms. Architectural distortion, a sign of advanced lung damage was seen in 37 patients (1%) in this series of studies. Most of these patients required ICU admission **(13)**. Nodules, including centrilobular nodules, solid nodules and tree-in-bud like densities, frequent findings in community-acquired cases of pneumonia were found to be less frequent in these studies. They were reported in only 81 cases (2%).

Three studies describe a new sign “spider web sign” – two of them by the same set of authors who report this sign in 25% of the patients with COVID-19 disease **(16,17)**. It is unclear whether there is any overlap of patients between these studies. This was originally described by Wu, J et al.as a triangular or angular GGO under the pleura with the internal interlobular septa thickened like a net causing retraction of the adjacent pleura in a spiderweb-like shape **(16)**. The only other study which reports this sign is by Zhu et al, they found this sign in 13%(4) of their patients **(18)**. In our opinion, this sign is very similar to the previously well known ‘pleural retraction sign’, which was described in two other studies included in the review **(12)**. Overall, the subpleural GGO or consolidation causing a web-like retraction of pleura was reported in 82 patients (3%). The same study by Zhou et al. **(12)** also reports ‘Vacuolar sign’ (a vacuole-like transparent shadow of < 5 mm in length observed in the lesion) in 54.8% (34 patients) of their study population. No other study included in this review describes this sign.

Sub-pleural linear opacities were reported overall in 2% (85) of the patients in these studies and sub- pleural transparent lines in 1% (33) patients.

Mediastinal lymphadenopathy and pleural effusion were less frequently found among the COVID-19 patients. They were reported in 40 (1%) patients and 66 patients (2%) overall. There is agreement among studies that there is higher incidence of pleural effusion and lymphadenopathy in critical patients and that they might reflect the viral load and virulence of COVID-19 disease (13, 17)

## 5. Discussion

### Terminologies used in the studies

Like in other radiology studies, these studies also used different terminologies as lesion descriptors. Such variations compromise the ability of researchers to derive meaningful insights from these studies. For example, only a few studies describe specific findings like vascular engorgement, “halo sign” or “reverse halo sign”. It is unclear whether these findings were not present in the other studies not describing these findings or the readers were not specifically asked to quantify these findings. Radiology Society of North America (RSNA) had acknowledged this pitfall and had released a consensus statement recommending the use of standardized language to reduce reporting variability. This also enables efficient data mining for future educational, research and quality improvement purposes.

In our reviewed studies, we encountered 49 descriptors for lung lesions. We mapped these descriptors to a shorter, unique list of 24 descriptors. This was done according to the Fleischner Society’s recommendations **(15)**. A recent study published in the New England Journal of Medicine surprisingly used X-ray descriptors like unilateral and bilateral shadowing to describe lesions on CT scans **(28)**. The list of these terminologies is provided in the supplementary material.

### Temporal change in the findings in the studies

During the early part of the outbreak, the hospitals in Wuhan, China were extensively using CT scans for screening patients with symptoms suggestive of novel coronavirus infection. Some studies showed better sensitivity for CT as compared to RT-PCR assay performed within 3 days of onset of symptoms **(19)**. The arguments in favor of such extensive use of CT were that RT-PCR tests of the sputum or nasopharyngeal swabs require several days with a false negative rate of more than 5% whereas CT imaging can show typical features of COVID-19 helping to rapidly screen and stratify patients **(6)**. Concerns were raised against such sweeping conclusions of the superiority of CT scans for the screening citing several reports that confirmed normal chest CT scan cannot exclude the diagnosis of COVID-19 **(20)**, especially for patients with early onset of symptoms. CT scans were normal in 11% to 56% of confirmed admitted cases during the early course of the diseases after the onset of symptoms but showed findings in the latter course of the disease **(21-23)**. But as the sensitivity of RT-PCR tests improved, the focus of studies shifted from using CT as a diagnostic tool to a prognostication test. A recent study from Italy **(10)** reported perilesional pulmonary vessel enlargement in areas of lung infiltrates could be an early predictor of lung impairment. Even though this finding was reported in several previous studies, no clear relationship to the prognosis of the patients is reported **(11-13)**.

### Temporal course of the disease

Most of these studies were cross-sectional studies, describing the CT findings at either initial presentation or during advanced symptoms. Among the 49 studies included in this review, there were only 14 studies, which included a total of 762 patients, which have described changes in the CT findings throughout the disease process. Chen et al., Wang et al. and Pan et al. reported the temporal changes in a series of 249, 90 and 63 patients respectively **(5, 7 & 9)**.

All these studies described a similar trend of changes. At the initial presentation, the common changes were patchy ground-glass densities or consolidations in sub-pleural lower lobe distributions **(Figure 3)**. Re-examinations scans done after 3-14 days showed progression and confluence of the GGO and consolidation leading to organizing pneumonia-like appearance in patients with worsening of symptoms **(Figure 4)**. Some patients showed intra-lobular septal thickening during this phase giving the classical “crazy-paving” appearance. If the initial scans showed nodules, the re-examination showed a reduction in the density of these nodules and merging of these nodules leading to a ground glass like density described as “melting sugar” appearance by Pan et al. Typically patients with a clinical improvement developed fibrotic stripes on these re-examination CT scans. In patients with further deterioration, needing ICU admission, the appearance was like a “white- lung” with diffuse high-density lesions in bilateral lungs.

**Figure 3.**
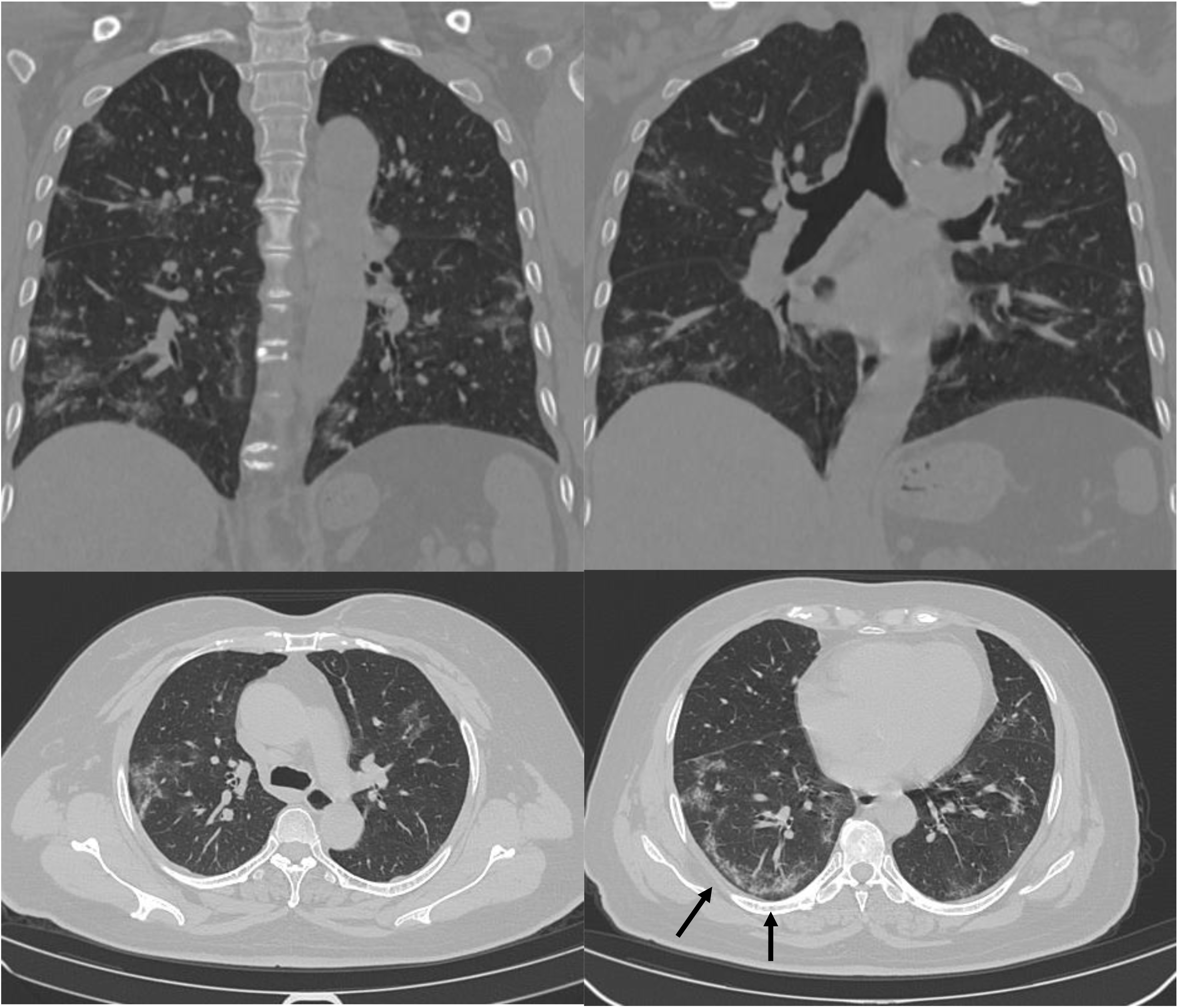
CT findings in a case of COVID-19 demonstrates diffuse bilateral ground-glass opacities with focal consolidations (mixed pattern) with reticulations in the lower lobes. Also note the sub-pleural sparing (arrow) seen in these lesions (image courtesy: www.coronacases.org)

**Figure 4.**
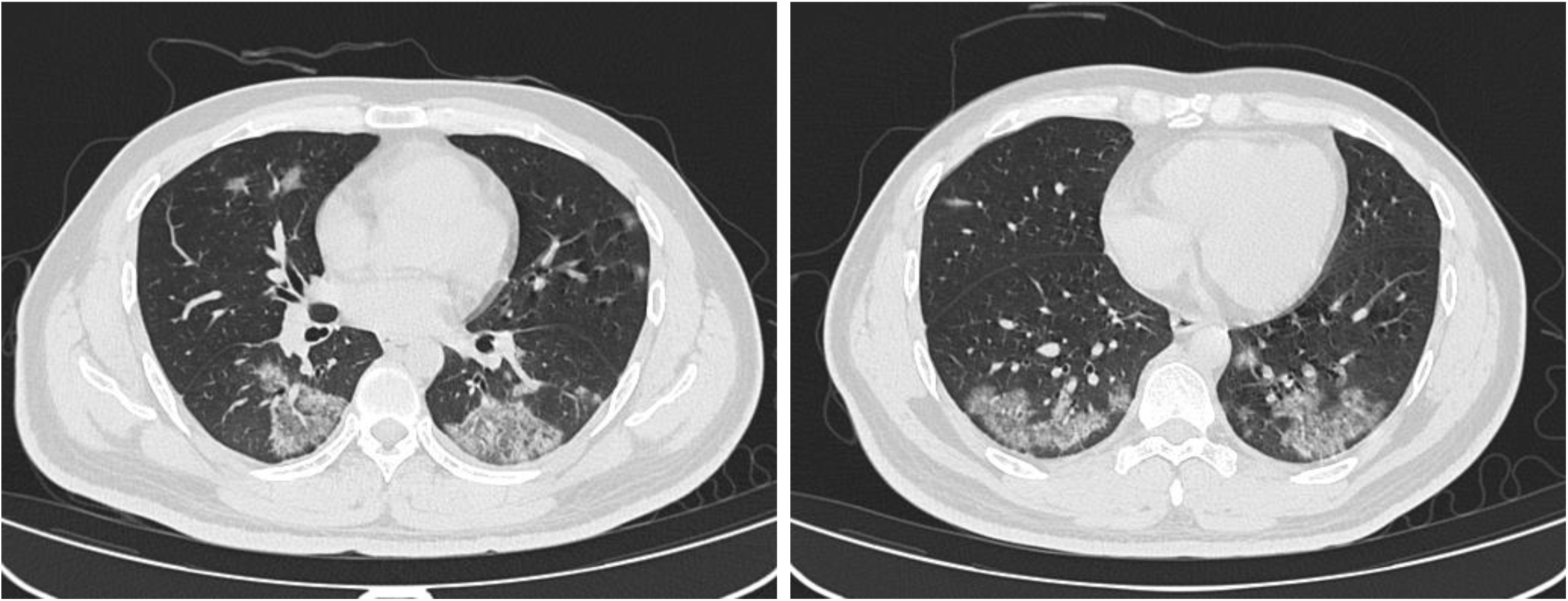
CT findings in a case of COVID-19 demonstrates bilateral peripheral ground-glass opacities with organizing consolidations (mixed pattern). There is also “reversed halo” appearance of central ground glass opacity surrounded by denser consolidation (image courtesy : www.coronacases.org)

### False-positive CT findings

CT findings of patients with negative results for RT-PCR tests were discussed only in 8 studies and included 530 (13%) patients out of 4145 patients. Considering only studies reporting negative results, the proportion of negative cases to positive cases was 34% (out of 1631 cases).

In the large series reported by Ai et al. of 1014 patients admitted in Wuhan **(4)**, there were 431 (41%) patients with negative RT-PCR results and 308 of these patients had positive chest CT scans (75%), while 97% (580/601) of the positive cases had positive chest CT Scans. All these patients were classified as having typical features of COVID-19 on CT scans. Such a high false positivity rate and low specificity (25%) can be explained by the fact that these cases were from the largest hospital in Wuhan, the central area of the outbreak during that period and many symptomatic patients with findings on chest CT were suspected to have COVID 19 disease to control the epidemic.

In another series reported by Zhu et al. out of 116 patients, 84 were negative for the presence of COVID-19 and 32 were positive **(18)**. Chest CT was performed in all these patients. 94% of the positive cases (30/32) and 67% of the negative cases (56/84) had changes of pneumonia on CT with bilateral involvement seen in 91% (29/32) and 40% (34/84) respectively. The most observed finding of GGO in other series were seen in only 47% of the cases in these series, whereas the more specific findings of a spider web pattern and crazy paving patterns were observed in even fewer patients, 13% (4 patients) and 3% (1 patient) respectively. GGO was observed in 12% of the negative cases also.

In a retrospective study by Cheng et al. that included 38 patients with clinical and epidemiological features consistent with COVID 19, 22 patients had negative RT-PCR findings for SARS-Cov-2 **(24)**. 11 patients had positive results. CT showed GGO in 90% (20/22) of the negative patients and all the positive patients whereas mixed GGO was observed in 72.7% (16/22) of the negative patients and 63.6% (7/11) of the positive cases. Consolidation was more frequently observed in negative cases (77.3%) as compared to 54.5% of positive cases. The etiology of the negative cases in the study remains unclear as these patients were rapidly discharged and could not be microbiologically confirmed. These studies demonstrate that the specificity of the most commonly reported CT findings of GGO & consolidation is too low to be able to use them as diagnostic features.

### Role of CT in initial false-negative RT-PCR cases

Fang et al. have reported better sensitivity of CT as compared to RT-PCR at initial patient presentation **(19)**. Among the 51 consecutive patients that they studied, only one had a normal CT scan at presentation. Of the 50 patients with abnormal CT scans at presentation, 36 (72%) had typical manifestations and 28% (14) had atypical manifestations. The initial RT-PCR test was positive in only 70% (36) patients, the second test is done in 1-2 days added 24% (12) more positive cases. Two more patients turned positive by the third test (2-5 days) and one patient showed positivity on the fourth test done 7 days after the initial onset of the symptoms. Even though the observations by the authors of better sensitivity at the initial presentation might seem right, the standard recommendation for declaring a patient negative for COVID-19 is the demonstration of negativity for SARS nCoV-2 genes in two consecutive tests done 24 hours apart. Following these criteria would have diagnosed 48 (94%) patients in this study within two days, even before the CT scan was performed. The average time from initial symptom onset to CT scan was 3 +/- 3 days in this study.

Similarly, in a study of 167 patients by Xie et al, there were 5 patients (3%) with negative RT-PCR results during initial tests who turned positive on repeat testing **(25)**. Bilateral multifocal GGO was observed in all these five patients. For a fair comparison, CT was negative in 7 RT-PCR positive patients (4%) at presentation.

Despite the conclusions by all these authors that CT is better for screening evaluation for COVID-19, critical evaluation of their results shows no clear evidence for the same.

### Pediatric patients have asymptomatic CT changes

Among the studies in this review, two studies discussed CT features of a total of 23 pediatric patients **(26, 27)**. Contact history and clinical symptoms of fever and cough were positive for all these patients. 7 patients had no findings on CT scans, 6 patients had unilateral and 10 patients’ bilateral lung field changes on CT scans. In the report by Xia et al. 60% of the pediatric patients had GGO and 50% had consolidation with positive ‘halo’ sign. 21 out of these 23 patients eventually recovered with two of them under observation until the time of the report. The initial findings and the longitudinal changes throughout the disease were found to be similar to adult patients with the initial patchy GGO’s and consolidations progressing to diffuse lesions with septal thickening, air bronchograms in the advanced stages, finally resolving completely or with residual fibrotic strips.

## 6. Conclusion

In this study, we have analyzed reports presenting CT features of 4145 patients from published literature in the PubMed database.

The lessons that the radiology findings of these studies offer are:

1. There is good agreement among these studies that diffuse bilateral GGOs is the most common finding at all stages of the disease.
2. There is a clear progression pathway of CT findings correlating with the clinical findings. Initial patchy GGO’s and consolidations progress to diffuse lesions with septal thickening, air bronchograms in the advanced stages. The further course can be either diffuse “white-out” lungs needing ICU admissions or complete resolution with or without residual fibrotic strips.
3. There is evidence that CT features like juxta- lesional pulmonary vascular prominence, pleural effusion & lymphadenopathy can be used as prognostic markers of COVID-19 disease.
4. We found no concrete evidence in our meta-analysis to favor the use of CT as an initial screening modality for COVID-19.
5. Wide variation in the lesion descriptors found in our metanalysis support the arguments in the statements made by radiology societies urging for use of standardized lexicons and structured reports. Importantly, we strongly recommend sharing of all imaging and report data that are the back bone of such research papers. This is more important during pandemics like COVID-19, since additional studies can be performed by researchers around the world to quickly develop tools and insights to fight off the disease.

## Data Availability

All the data files related to this study are provided as supplementary material.

## Footnotes

1 Readers of the studies

